# COLLABORATIVE MAPPING AS A METHODOLOGY FOR IDENTIFYING COMMUNITY PERCEPTIONS ON BASIC SANITATION NEEDS AND INTERVENTIONS FOR LEPTOSPIROSIS IN SALVADOR, BRAZIL

**DOI:** 10.64898/2026.03.06.26347767

**Authors:** Fabiana Almerinda G. Palma, Pablo Ruiz Cuenca, Daiana Santos de Oliveira, Ana Maria N. Silva, Yeimi Alexandra Alzate Lòpez, Diogo César de C. Santiago, Marbrisa N. R. das Virgens, Ariane Sousa do Carmo, Antonia dos Reis, Gislane de Jesus do Carmo, Andrea Maria Lima, Renata Santos Almeida, Lucineide Oliva, Juliet O. Santana, Pedro Maciel, Tania Bourouphael, Emanuele Giorgi, Ricardo Lustosa, Max T. Eyre, Caio G. Zeppelini, Cleber Cremonese, Federico Costa

## Abstract

Despite the relevance of spatial mapping in analyzing the health situation and understanding the risk factors and determinants of leptospirosis, peripheral urban communities often remain invisible on maps, which tend to use data and methods that do not express community contribution nor promote local participation. Furthermore, in the implementation of sanitation interventions, the same happens: there is limited user participation, and a lack of identification of intervention needs based on the perception of community residents, failing the interventions. We conducted a cross-sectional study through collaborative mapping from February to October 2022 with 213 residents and self-declared heads-of-household in two peripheral urban communities. We analyzed the perception of sanitation needs indicated by residents and their relationship with the risk of leptospirosis in these communities. Based on community perception, sewage (NS: 87.1%; JSI/ME: 84.9%) and urban cleaning and solid waste management (NS: 25.9%; JSI/ME: 32.6%) were the sanitation needs. In NS, most participants indicated that the necessary interventions for sewage improvement were actions of sewer cleaning and sealing (26.5%), sewer cleaning and piping (23.5%), and implementation/installation/construction of a sanitary sewage network (41.4%). In JSI/ME, interventions included sewage sealing (48.7%) and piping (25.6%), in addition to actions to maintain sewage cleaning (93.3%). The removal of solid waste (trash) in the square (NS: 22.2%) and on the streets (JSI/ME: 69.2%), as well as community awareness (JSI/ME: 15.4%), were indicated as interventions to meet the needs of urban cleaning and solid waste management. Respondents agreed on where interventions should occur, which congregated around the local river. We found a negative correlation between the predicted leptospirosis seropositivity and perceived intervention needs in both study areas. The prevention of diseases such as leptospirosis in peripheral urban communities requires integrated basic sanitation interventions, encompassing different components and aligned with the local needs perceived by residents.

## Introduction

In low-and middle-income countries (LMICs), zoonotic diseases remain a public health problem. Among these diseases, urban leptospirosis affects populations in situations of greater social vulnerability, who reside in territories with precarious sanitation and infrastructure conditions [1–5]. Leptospirosis is a neglected disease [6] caused by pathogenic spirochete bacteria of the genus *Leptospira* [4,5]. Its main reservoir in urban areas is *Rattus norvegicus*, which sheds pathogenic leptospires into the environment through urine [6,7].

Knowledge of the environment favors the understanding of the distribution of leptospirosis, the living conditions of people [8], as well as the main determinants of disease [9,10]. Furthermore, it is an essential resource for identifying the unequal impact of the main risk factors that contribute to the maintenance of these diseases in socially vulnerable contexts [10]. This knowledge also makes it possible to identify determinants, such as sanitation, which play a crucial role in maintaining leptospirosis [1].

The use of participatory methodologies, such as collaborative mapping, for environmental knowledge of leptospirosis facilitates the identification of determinants of and risks to health, allowing the proposal of more effective interventions to prevent these risks [11]. This methodology can favor the implementation of timely interventions and ensure greater sustainability of actions through the participation of communities, especially in areas marked by extreme vulnerabilities and historical, environmental and social inequalities [12,13].

Collaborative mapping is a map creation process in which the population voluntarily contributes by generating relevant content [14], actively participating in the process of recognizing the territory in which they live [15] through the use of integrated methods and technologies [16]. This approach is considered a valuable instrument for popular participation [17], as it allows local social actors to fill gaps in official maps [18], build a solid understanding of the resources, health problems, risk factors and determinants of the diseases that affect them [19]. In addition, it promotes the identification of future interventions aimed at reducing or eliminating problems that pose a risk to life.

Diagnosing health risks and determinants through special analyses that use participatory methodologies is essential in analyzing the health situation. It is a key factor in understanding the necessary interventions, especially in places with absent or insufficient basic sanitation services, such as sewage and urban cleaning [20]. In contexts with problems related to sewage, a determinant of leptospirosis, it is common to observe limited community participation in supporting the construction of projects and the management of construction. Community participation is often limited to specific agreements allowing pipes to pass through private properties [21], which may be one of the factors contributing to the persistence of local problems related to these services.

Although there are official maps containing information on risk factors and determinants in territories that suffer from vulnerability processes [22], some areas of these environments often remain invisible or do not have updated information on the maps [23]. Existing maps lack adequate information on hard-to-reach areas [24] and predominantly use techniques that do not encourage community participation. Traditional mapping tools and techniques such as Geographic Information Systems (GIS) and geomorphological mapping [25] are not capable of meeting the needs of local communities due to their objectives and scales. Furthermore, excessive control and lack of communication between public agencies responsible for collecting data and generating information often result in the lack of spatial data related to infrastructure and socioeconomic conditions at the municipal level [24,25].

The lack of detailed information about the community in urban environments hinders responses to public health emergencies and disasters [24], as well as a broader understanding of risk factors and determinants of health. It also restricts the planning of interventions to priority areas, as perceived by communities. This issue is particularly relevant in interventions related to sanitation [24] and the prevention of diseases such as leptospirosis, which continue to affect populations in low-income communities in large urban centers. Previous studies have reported that poor environmental and sanitation conditions increase the risk of diseases such as leptospirosis [1,9,10]. However, few studies have analyzed residents’ perceptions of the need for sanitation and the solutions and interventions suggested by those living in contexts with these deficiencies. In this study, our objective is to collaboratively identify the need for interventions in basic sanitation as perceived by residents, their spatial distribution, and their relationship with leptospirosis seropositivity in two communities where sanitation interventions were implemented.

### Ethics Statement

This study was approved by the Research Ethics Committee of the Institute of Collective Health/Federal University of Bahia (CEP/ISC/UFBA) with CAAE number 32361820.7.0000.5030, and a National Research Ethics Committee (CONEP) linked to Brazilian Ministry of Health under approval number 4.235.251. All participants were informed about the study and gave their written consent to participate.

## Materials and Methods

### Study characteristics

This study is a cross-sectional study that is part of a participatory research project, which integrates space components on spatial community diagnosis of vulnerabilities associated with sanitation to improve health and reduce health disparities. The initial research was conceptualized as an expansion of the objectives of a previous study [26] aimed to analyze the perception of environmental health risks through collaborative mapping, focusing on the factors contributing to leptospirosis in four peripheral communities of Salvador, Brazil. This study was carried out by the research group and through a collaboration between people from the communities and academic researchers with study experience using collaborative mapping in similar communities. Thus, the core research team comprised community researchers (people who lived in the communities at different sites where the study was conducted), and academic researchers. These acted as supporters of the communities’ residents and contributed to field research and data analysis.

### Study Area

This study was part of a research project on health intervention and prevention of urban leptospirosis, which was started in 2021 in communities in Salvador, Brazil [27]. In these communities, we carried out a baseline serological survey with the application of individual and household questionnaires, and collection of biological samples collected by trained phlebotomists. In addition, we conducted community engagement in two areas for the involvement and participation of residents in research project activities [27].

This study integrates one of the components of the community engagement model from the research project, collaborative mapping, which enabled extensive spatial data collection in the communities. The study was carried out in the neighborhoods of Nova Sussuarana (NS) and Jardim Santo Inácio/Mata Escura (JSI/ME). These neighborhoods are located in the periphery of the city Salvador [28], Brazil (Fig 1), and have a population which is mostly low-income, self-declared black, and with sanitation problems [1,29]. Unadjusted seroprevalence for *Leptospira* in NS and JSI/ME in the baseline survey was 9.0% (n= 338) and 8.0% (n= 205), respectively.

**Fig 1.**
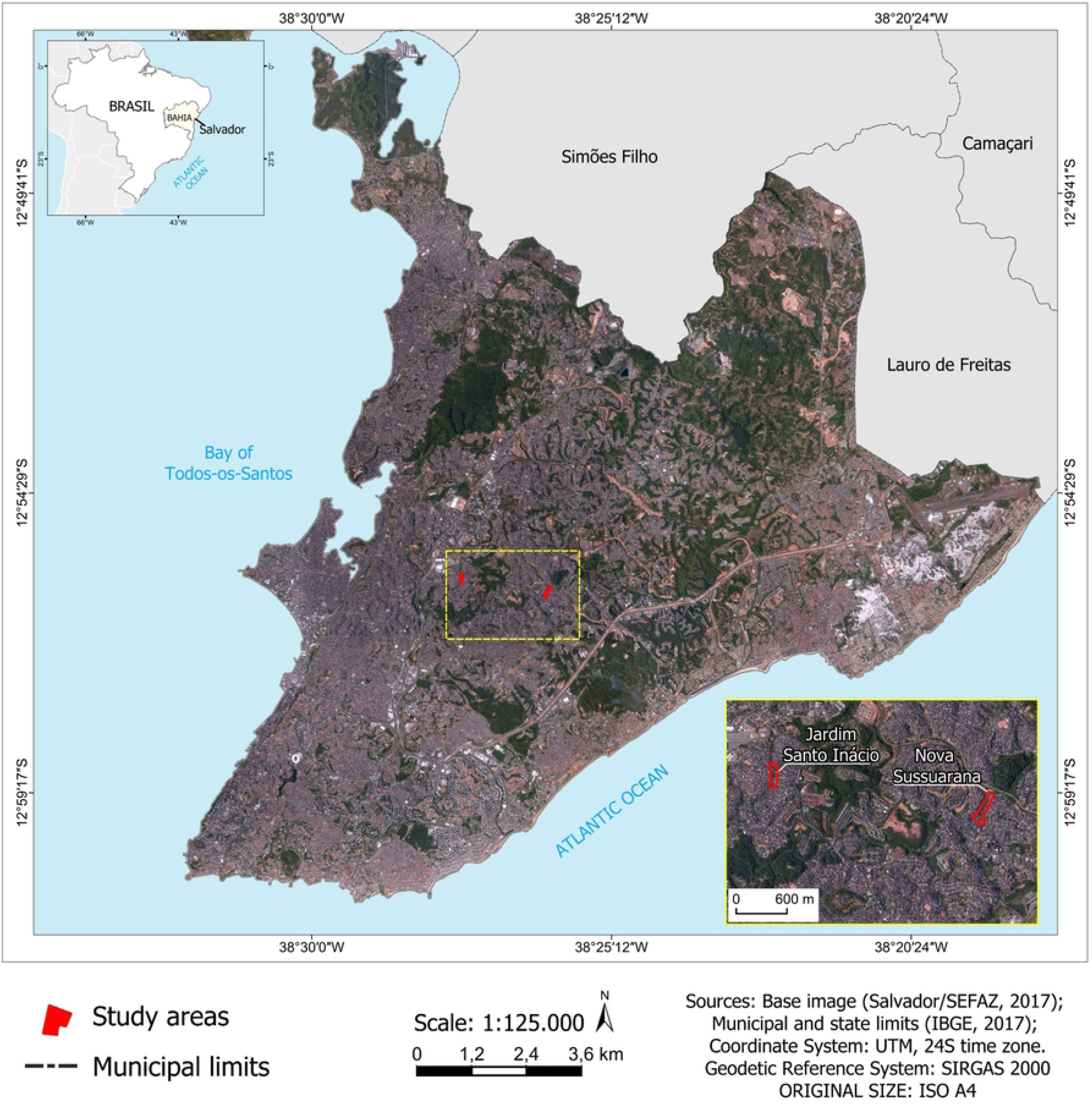
Map of the two study areas within the city of Salvador. The map in Figure 1 was created by the authors with open secondary data produced by the authors using the free and open-source software QGIS, version 3.38, available for download at: https://www.qgis.org/pt_BR/site/forusers/download.html. The map data contains municipal, state, and country boundaries provided by the Brazilian Institute of Geography and Statistics (IBGE, 2017; 2021), Coordinate System: UTM, 24S time zone, and Geodetic Reference System: SIRGAS 2000. The open-access aerial photography is also available on the Salvador City Hall website (Salvador, 2017). All mentioned data can be consulted with their respective metadata on the websites: https://www.ibge.gov.br/geociencias/organizacao-do-territorio/malhas-territoriais/15774-malhas.html?=&t=downloads; https://www.ibge.gov.br/geociencias/organizacao-do-territorio/malhas-territoriais/15774-malhas.html; http://mapeamento.salvador.ba.gov.br/geo/desktop/index.html#on=layer/default;bairros/bairros;scalebar_meters/scalebar_m;orto2016/Ortoimagem_Salvador_2016_2017&loc=76.43702828517625;-4278080;-1445884.

### Study population

In this study, we used a census sample from the serological survey carried out in the study areas and approached those eligible to participate in this collaborative mapping study in NS (N=124) and JSI/ME (N=121). Eligible residents were (1) men and women self-declared heads-of-household (if these were absent after three consecutive home visits by the team, any family member aged 18 or older); (2) who slept in the household for at least three nights per week; (3) had participated in the serological survey in 2021; and (4) were able to provide written informed consent.

### Data collection

#### Collaborative mapping

In our study, people who live in the communities were involved in all phases of the collaborative mapping, and the research process itself had a goal of social action related to using the study results to claim improvements in sanitation and infrastructure in their communities. Members of the research team and residents of Nova Sussuarana (NS) and Jardim Santo Inácio/Mata Escura (JSI/ME) participated in virtual and face-to-face training.

Due to the COVID-19 pandemic, the virtual meeting was planned for a maximum total of 10 team members, where eight were residents of the communities that actively participated in the construction of the training proposal, and the following topics were worked on: 1) Collaborative mapping: concepts, methodology, and results of the previous research project; 2) Dialogued exposition about a territorial reading of the city of Salvador and its socio-spatial segregation; 3) Social determinants of health and zoonosis; 4) Maps: definitions and uses, and location of the study area; 5) Manipulation of web map projects in the Vicon Saga system [30]; 6) Data migration from physical maps to QGIS and its processing.

After the training, the team visited participating residents between February and March 2022, for those in NS and between August and October for JSI/ME, and explained the research objectives before inviting them to participate. The team used a standardized questionnaire to capture the participants’ perceptions of basic sanitation needs, what potential solutions they would recommend, and where these should be implemented, giving them sufficient time to consider their answers. At this stage, resident participation occurred at the individual consultation level. This questionnaire was validated in a previous study carried out in a community with similar characteristics to those included in this study which is awaiting publication. The research team used printed maps to help participants locate themselves, highlighting local landmarks. The participants were then able to draw their perceptions on a separate map. Data on the type of basic sanitation that required intervention were based on the classification proposed in Brazilian Law for sanitation [31].

At the same time, we classified the data on the types of intervention for basic sanitation reported by the participants into the following dimensions: implementation/installation/construction, maintenance, removal, integrated action, and others. These were based on free text answers given by participants. We categorized the data through thematic analysis [32] and subsequently validated it by a specialist in the field of basic sanitation. Implementation/installation/construction interventions corresponded to works that had not been carried out previously in the communities. Interventions classified as maintenance corresponded to improvements, suggested by participants, in sanitation works carried out in the past. Removal interventions involved removing garbage from specific locations in the communities. Those classified as integrated actions corresponded to reported interventions that comprised more than one of the mentioned components of basic sanitation. For this study, we considered river and sewage as synonymous and garbage basket and garbage containers are treated as having the same meaning as informed by participants during the research. In the case of sewage, due to the pollution of water bodies by domestic garbage and the deficiencies in the sewage system, residents began to perceive the river as part of an open sewer.

Our questionnaire was organized into six parts: 1) Health risk indicated by residents (open field variable) and the level of risk for the problem indicated (Low/Medium/High); 2) Coverage of primary care health services; 3) Indication of the health risk for the resident on the map; 4) Health promotion locations in the neighborhood (the type of location and indication on the map); 5) Intervention and perception on basic sanitation; 6) Use of cell phones by residents: Do you have a cell phone (Yes/No), Type of cell phone (Smartphone/Basic cell phone).

For this study, we worked with questions regarding the perception of basic sanitation needs and the types of interventions suggested by the participants. Here, the variables that worked on the perception of basic sanitation were: “Need for intervention close to the residence (yes/no)”; “Type of basic sanitation intervention (water supply, sewage, urban cleaning and solid waste management, urban drainage and rainwater management, others)”; “The type of intervention to solve the problem (open question),” which was categorized into five dimensions: implementation/installation/construction; maintenance; removal; integrated action; others.

Given the number of pieces of information that could be identified on the physical map regarding the sanitation intervention indication, the field team used a code and the following strategy presented in data collection: recording points, lines, or polygons which residents used to indicate the need for carrying out interventions on the components of sanitation, with the letters representing the initials of the components in Portuguese: i) sewage (E); ii) water supply (AA); urban cleaning and management of solid waste (LU); iii) urban drainage and stormwater management (D); iv) others. In this study, the “ID Vértice” served as an identification key for each problem/phenomenon indicated by the resident, which was represented through the georeferencing of geometric primitives (point, line, and polygon), using GIS for creating maps. At this stage, resident participation occurred by systematizing data on the map and discussing results in collective meetings.

The maps in A4 format were prepared to facilitate visualization by the residents during data collection. Each printed map also had a space for filling in the identification number given by the project for the household and a small caption containing the numbers related to each item identified on the maps.

#### Serological data

Serological data to identify previous exposure to *Leptospira* was collected by trained phlebotomists after written participant consent. Serological analyses were conducted using the Microscopic Agglutination Test (MAT), as described in Khalil et al. [1] and De Oliveira et al. [33]. The Microscopic Agglutination Test (MAT) is the gold-standard diagnostic method for leptospirosis [34]. We used a diagnostic panel that included the most prevalent serovars in the region as well as other known pathogenic serovars. These were *L. kirschneri* serovar Cynopteri strain 3522C, *L. kirschneri* serovar Grippothyphosa strain Duyster, *L. interrogans* serovar Canicola strain H. Ultrech, *L. interrogans* serovar Autumnlais strain Akiyami A, *L. borgspetersenii* serovar Ballum strain MUS 127, *L. interrogans* serovar Copenhageni strain Fiocruz L1-130 and *L. interrogans* serovar Copenhageni strain Fiocruz LV3954. Seropositivity was defined as ≥50% agglutination of leptospires at a titer of ≥1:50. Samples reactive at a 1:100 dilution were further titrated in serial dilutions. The presumptive infecting serogroup was considered the one with the highest titer, at least one dilution above the others; Titers <1:50 were considered negative [1].

#### Statistical analysis

Descriptive analyses were carried out to show differences within and between study areas, using STATA Statistical Software version 14 [35]. To assess sociodemographic, household, peri-domiciliary, and perceived sanitation needs variables, see Tables 1, 2, and 3 and supplementary materials S1, S2, and S3.

**Table 1.** Sociodemographic characteristics of the study population.

| Variables |  | Overall<br>(N= 212) | Communities |  | p-value |
| --- | --- | --- | --- | --- | --- |
|  |  |  | NS* | JSI/ME** |  |

|  | Responses |  | (N=107) | (N= 105) |  |
| --- | --- | --- | --- | --- | --- |
|  |  | Frequency (%) |  |  |  |
| **Sex** | 212 |  |  |  |  |
| Female |  | 154 (72.6) | 76 (71.0) | 78 (74.3) | 0.595*** |
| Male |  | 58 (27.4) | 31 (29.0) | 27 (25.7) |  |
| **Age (years)** | 211 |  | **N= 106** | **N= 105** |  |
| ≤ 40 |  | 109 (51.7) | 56 (52.8) | 53 (50.5) | 0.732*** |
| > 40 |  | 102 (48.3) | 50 (47.2) | 52 (49.5) |  |
| **Ethnicity <sup>1</sup>** |  |  |  |  |  |
| White |  | 8 (3.8) | 4 (3.8) | 4 (3.8) | 0.089**** |
| Black |  | 109 (51.7) | 56 (52.8) | 53 (50.5) |  |
| Mixed |  | 88 (41.7) | 46 (43.4) | 42 (40.0) |  |
| **Education** | 167 |  | **N= 85** | **N= 82** |  |
| ≤ Primary school |  | 77 (46.1) | 37 (43.5) | 40 (48.8) | 0.496*** |
| ≥ Secondary school |  | 90 (53.9) | 48 (56.5) | 42 (51.2) |  |
| **Occupation** | 100 |  | **N= 54** | **N= 46** |  |
| Informal work |  | 57 (57.0) | 32 (59.3) | 25 (54.3) | 0.621*** |
| Formal work |  | 43 (43.0) | 22 (40.7) | 21 (45.7) |  |
| **Government assistance <sup>2</sup>** | 208 |  | **N= 103** | **N= 105** |  |
| No |  | 157 (75.5) | 56 (54.4) | 101 (96.2) | <0.001**** |
| Yes |  | 51 (24.5) | 47 (45.6) | 4 (3.8) |  |

\*NS: Nova Sussuarana
\*\*JSI/ME: Jardim Santo Inácio/Mata Escura
<sup>1</sup> 6 (5.7%) participants self-declared of Asian ethnicity (JSI/ME community)
<sup>2</sup> Bolsa Família or Benefício de Prestação Continuada
\*\*\* Chi-square test
\*\*\*\* Fisher's exact test

**Table 2.**
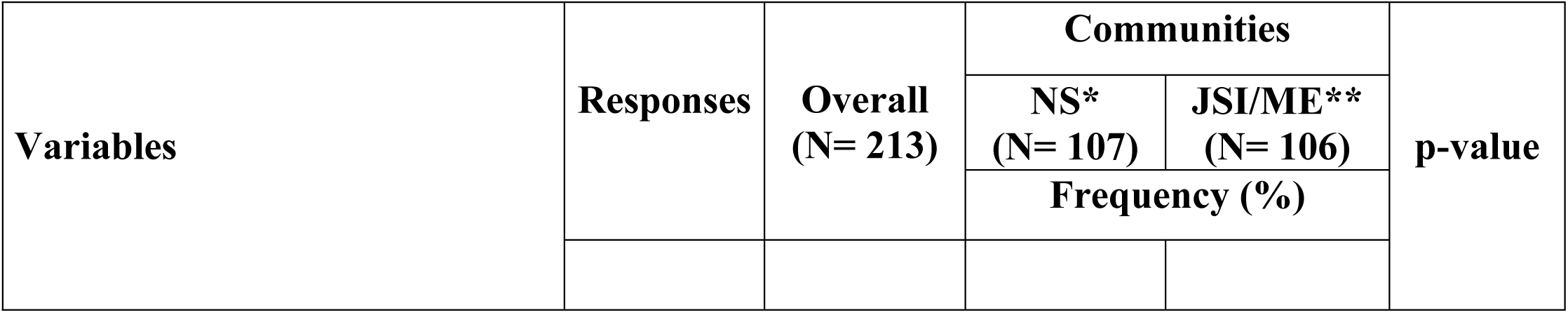
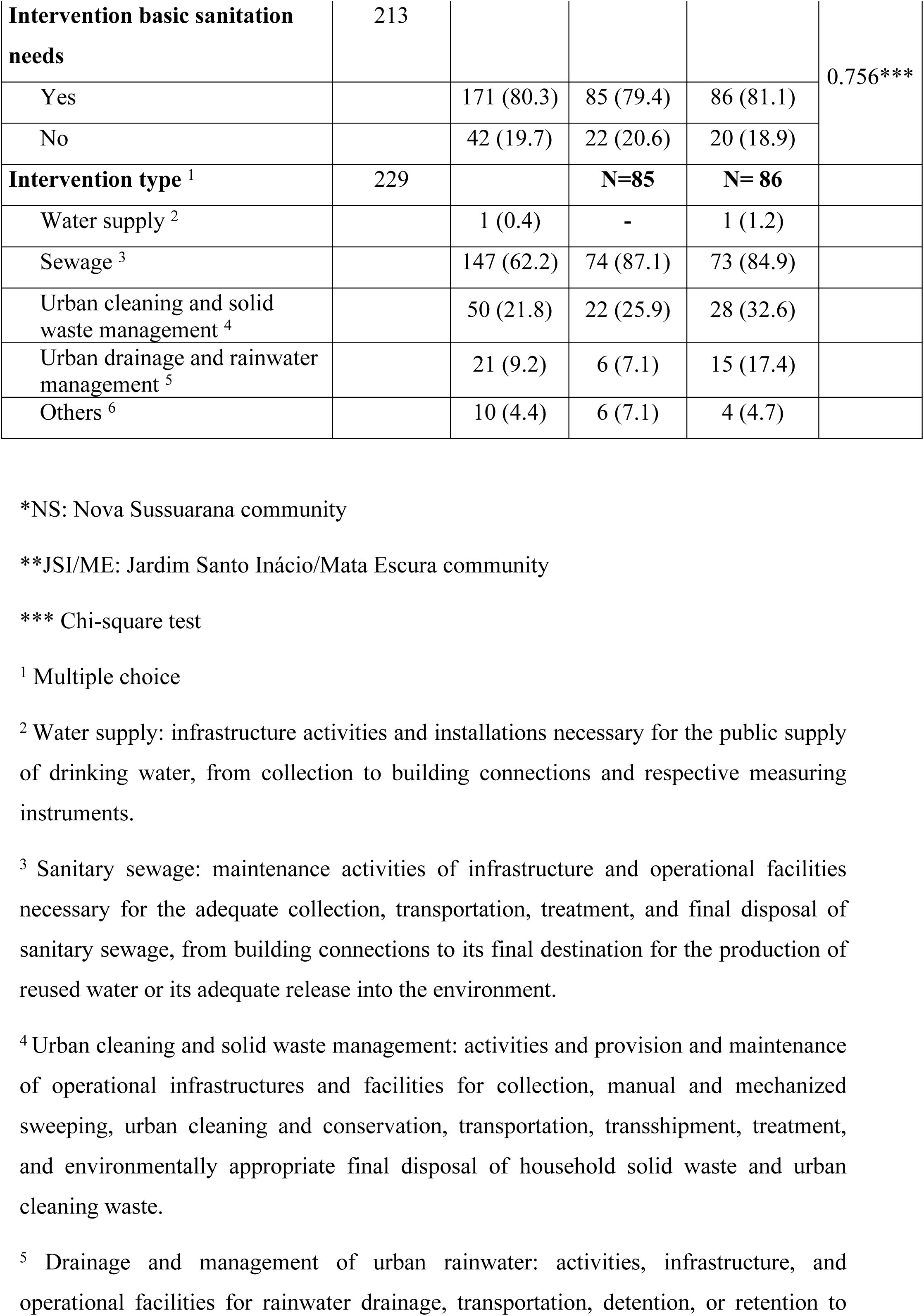
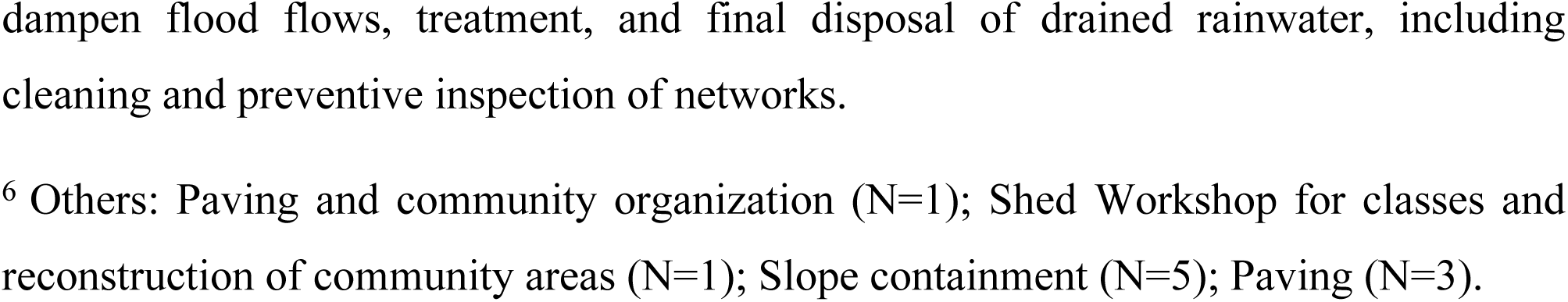
Perceived and type sanitation needs reported by the study population.

##### Seropositivity model

To assess the correlation between leptospirosis seropositivity, used as a proxy for leptospirosis risk, and perceived intervention needs, we used a logistic regression model to predict seropositivity across each study area. These analyses were carried out in R, version 4.2.1 [36]. For this, we used individual level data, including the serological data described above, from residents across both study areas. Individuals had to be living in one of the study areas for at least the last 6 months, sleep there at least 3 nights a week, be at least 18 years old and be able to provide written consent. Each individual also answered a questionnaire that collected their age and gender, amongst other characteristics. We also recorded the location of their households.

A team of researchers visited the study areas to record the locations of a number of spatial features, including known and possible risk factors (supplementary materials S6, S9 a S11). Minimum Euclidean distances from individuals’ households and each of these features were calculated. Alongside these features, we created a land cover raster of each study area using satellite images taken by WorldView-3, with a resolution of 0.3 meters. These rasters had three mutually exclusive land cover classes: soil, vegetation and impervious land. A 20-meter circular buffer was created around each household to calculate the percentage of each land cover class within this area. In the analysis, each individual was linked to their household’s data.

To create the logistic regression model, we started by exploring the associations between all these spatial variables and individual leptospirosis serological status, as binary variable. We used a binomial generalised additive model (GAM) for the exploratory phase [37], which included age, gender and study area as a priori variables [10]. These models gave us the possibility to assess whether the variables should be included as linear or non-linear relationships. Multi-variable models were built using the Akaike Information Criterion (AIC), selecting the lowest value. Variables were grouped by themes, such as land cover or roads (see Supplementary Material S6 for full list of groups). Priority was given to models where a whole group was removed. More detailed information is available in the supplementary material S6.

The final model included the distance from households to pruning rubbish piles, the percentage of soil land cover within a 20-meter circular buffer, as a non-linear relationship with a knot at 25%, and the *a priori* variables mentioned above (age, gender and study area). Model coefficients were applied across raster values for these variables, keeping age and gender fixed as 35 year old male, to predict the probability of leptospirosis seropositivity for every cell in a predictive raster. The demographic characteristics had to be maintained constant to create static layers for maps. We chose these specific characteristics as they are considered the highest at-risk group for leptospirosis infection.

##### Perceptions vs. predictions

The perception data were available as georeferenced vector data, in either point, line or polygon form. To standardise the data, all points and lines were given a 5-meter buffer to create polygons. A grid with a resolution of 5 meters was laid over each study area. For all points on the grid, the density of each kind of perceived intervention need was calculated, using the perception polygons. As there was only one polygon for perceived intervention on water supply, this was excluded from further analyses. The predicted leptospirosis seropositivity was also extracted for each of these points (Fig 2).

**Fig 2.**
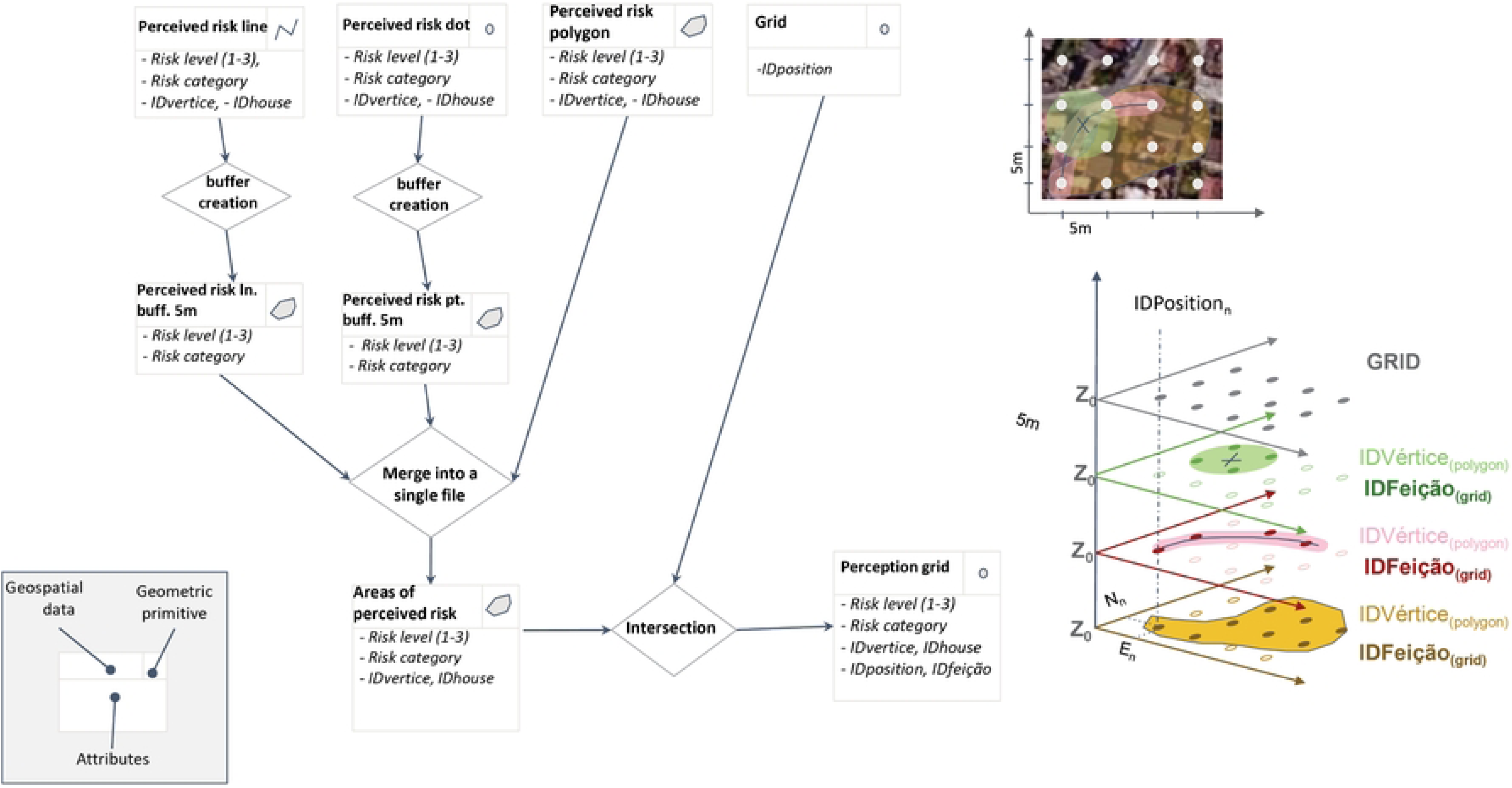
Diagram to show how perception data was standardised into a grid

A Poisson regression model was fitted to the predicted leptospirosis seropositivity and the densities of perceived intervention need, both of which had values for each grid point in the studies areas. The model estimated how predicted seropositivity, the exploratory variable, increases the rate (or density) of perceived intervention need identified by the participating residents, the response variable. Here, model coefficients quantified the associations between objective values (predicted seropositivity) and perceived values (perceived intervention need). A separate model was used for each of the intervention needs: drainage, sewage system and urban cleaning.

Given that we had the individual characteristics of the participants who supplied perception data, we were also able to perform a subgroup analysis looking at how these effects changed with gender and leptospirosis serological status.

#### Ethics statement

This study was approved by the Research Ethics Committee of the Institute of Collective Health/Federal University of Bahia (CEP/ISC/UFBA) with CAAE number 32361820.7.0000.5030, and a National Research Ethics Committee (CONEP) linked to Brazilian Ministry of Health under approval number 4.235.251.

## Results

### Sociodemographic characteristics

Among 245 eligible residents from the communities (NS= 124; JSI/ME= 121), 213 (86.9%) were enrolled in this study (NS= 107, 50.5%; JSI/ME= 106, 49.5%), with a response rate greater than 80% in each of the communities. There was homogeneity for most of the participants’ sociodemographic characteristics in the study communities, except for receipt of government assistance. Most participants who received government assistance, Bolsa Família or Benefício de Prestação Continuada, lived in NS compared to JSI/ME (44.8% vs. 3.8%; p-value<0.001) (Table 1).

### Households’ and peri-domiciliary characteristics

Household locations are shown in Figure 3. Regarding peri-domiciliary characteristics, in NS, more households had cement as the primary flooring material (17.8% vs. 6.7%; p-value=0.008) and were close to slopes (26.2% vs. 10.4%; p-value=0.003) compared to the household of participants from JSI/ME. In JSI/ME, more households did not have a Water meter (64.6% vs. 48.6%; p-value=0.024) compared to households in NS. Furthermore, a higher percentage of participants living in JSI/ME also reported more problems with the lack of water in their homes, the equivalent of 3 to 6 times a week (25.3% vs. 3.7%; p-value<0.001) compared to households in NS. In NS, a higher percentage of households did not have paved access to the house (15.9% vs. 3.8%; p-value=0.005) compared to households in JSI/ME (S1 Table).

**Fig 3.**
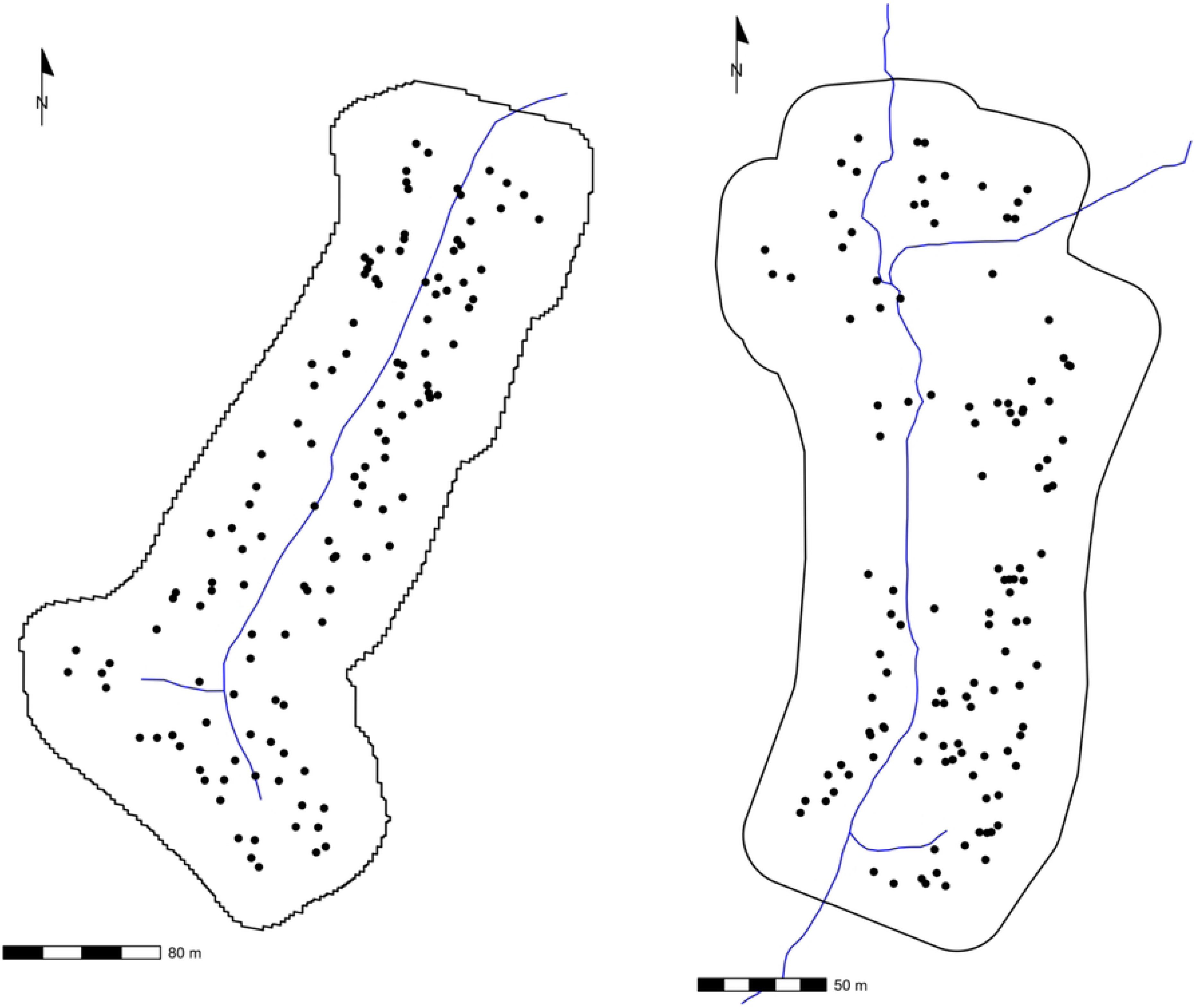
Household locations jittered randomly by 5 meters to maintain anonymity. The authors created the map in Figures 3 using the results generated from their primary data analysis.

### Perceived basic sanitation needs and intervention

There was no statistically significant difference in the perception of the need for basic sanitation intervention between participants who lived in NS and JSI/ME. Furthermore, sewage (87.1% NS vs. 84.9% JSI/ME) was the most reported intervention for participants in the two study communities, followed by urban cleaning and solid waste management (NS: 25.9%; JSI/ME: 32.6%). Despite that, a higher percentage of JSI/ME participants also reported the need for urban drainage and stormwater management intervention (17.4% vs. 7.1%; p-value=0.037) compared to the participants in NS (Table 2).

After selecting key perceived problems of basic sanitation, participants were asked to suggest potential interventions for the indicated needs. All responses were grouped into four categories (Table 3, and supplementary materials S2, S3), followed by the types of interventions indicated for each. When it came to sewage, the NS participants pointed out a variety of interventions. Integrated action (44.6%) and implantation/installation/construction (39.2%) were the most reported, with different measures suggested for each category. For instance, integrated sewage actions were proposed for sewer cleaning and sealing (26.5%), sewer cleaning and piping (23.5%).

**Table 3.**
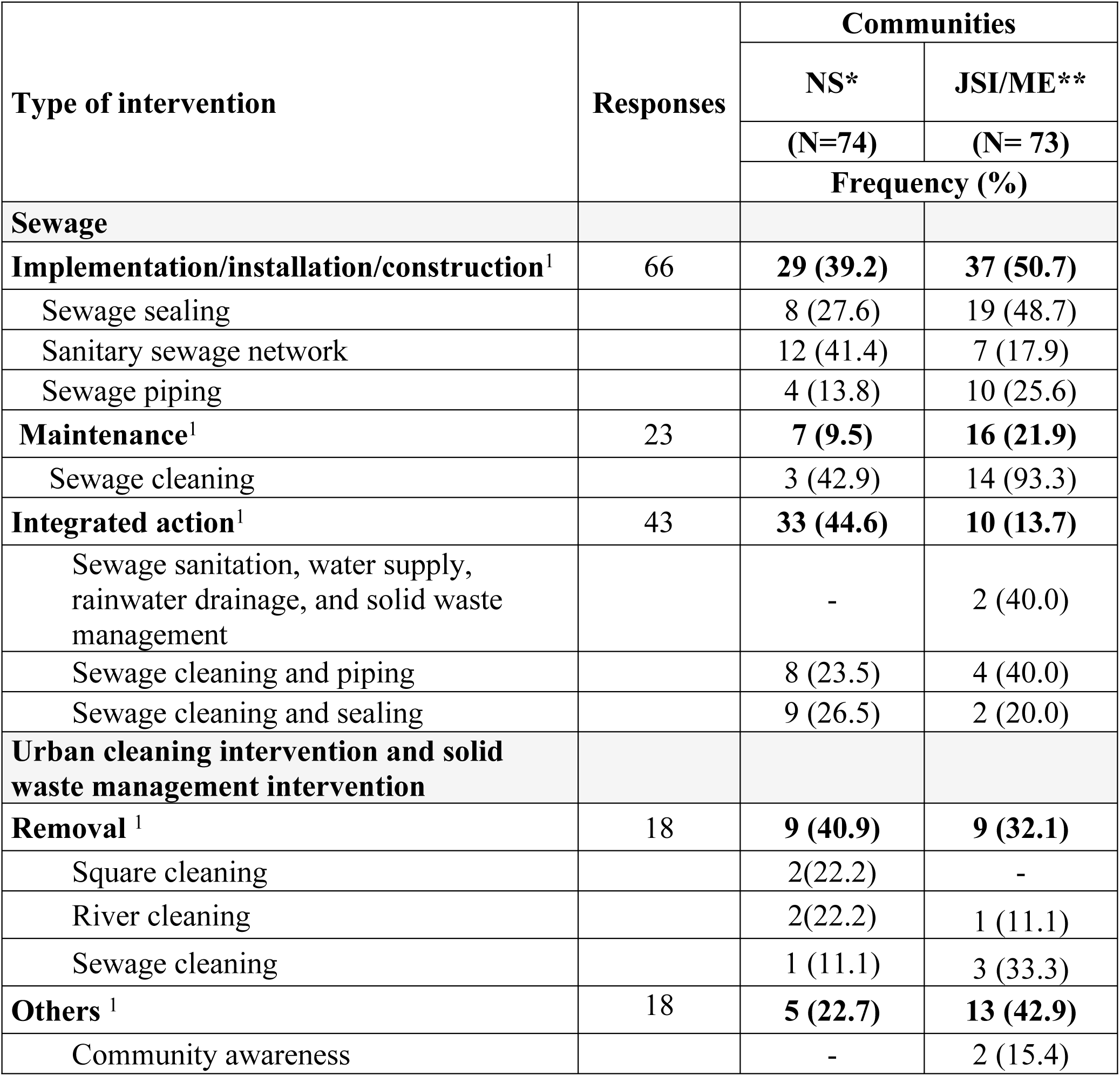
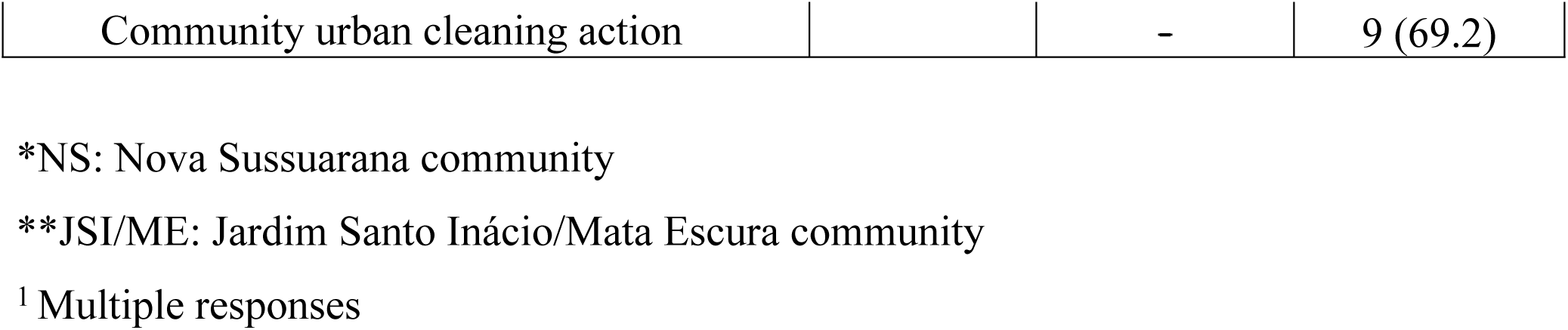
Type of sewage and urban cleaning and solid waste management interventions reported by the study population.

For implementation/installation/construction interventions, the implementation of the sanitary sewage network and sewage piping (41.4%), in addition to sewer sealing (27.6%), was the most indicated by study participants (Table 5). In JSI/ME, most interventions indicated by participants were related mainly to implementation/installation/construction (50.7%) and maintenance (21.9%) of sewage systems dimensions. For implementation/installation/construction interventions were reported o sewage sealing (48.7%) and sewage pipe (25.6%). Thus, most indications for sewage maintenance interventions were cleaning (93.3%) (Table 5). See the supplementary material (S2 Table) for a full table of the type of sewage interventions reported by the study population.

For urban cleaning and solid waste management interventions, most participants in study communities proposed measures in different categories, with interventions related mainly to trash removal (40.9%) in NS and others (42.9%) in JSI/ME. In the trash removal category, the most reported interventions in NS were mainly to clean the square (22.2%) and river (22.2%), while in JSI/ME, in the category others, cleaning the community streets (69.2%) and community urban cleaning action (69.2%) were the most mentioned by participants (Table 3). See the supplementary material (S3 Table) for a full table of the type of urban cleaning and solid waste management interventions reported by the study population.

### Relationships of perceived basic sanitation intervention needs and the risk of infection from leptospirosis

The probability of leptospirosis seropositivity in a 35-year-old male was unequal throughout both study areas (Fig 2). There were clear hot-spots located across the study areas, with a probability of up to 49.9% in NS and 44.3% in JSI/ME.

The distribution of perceived intervention needs also varied across the study areas (Fig 4). The perceptions for all three intervention types clustered around the local river in both communities. The densities varied greatly: the highest density was for sewage interventions, with a maximum of 67 perceptions per point in NS and 54 in JSI/ME; the second highest was for urban cleaning, with a maximum density of 7 in NS and 12 in JSI/ME; and the lowest was drainage, with 4 perceptions per point in NS and 5 in JSI/ME as maximum densities (Fig 4).

**Fig 4.**
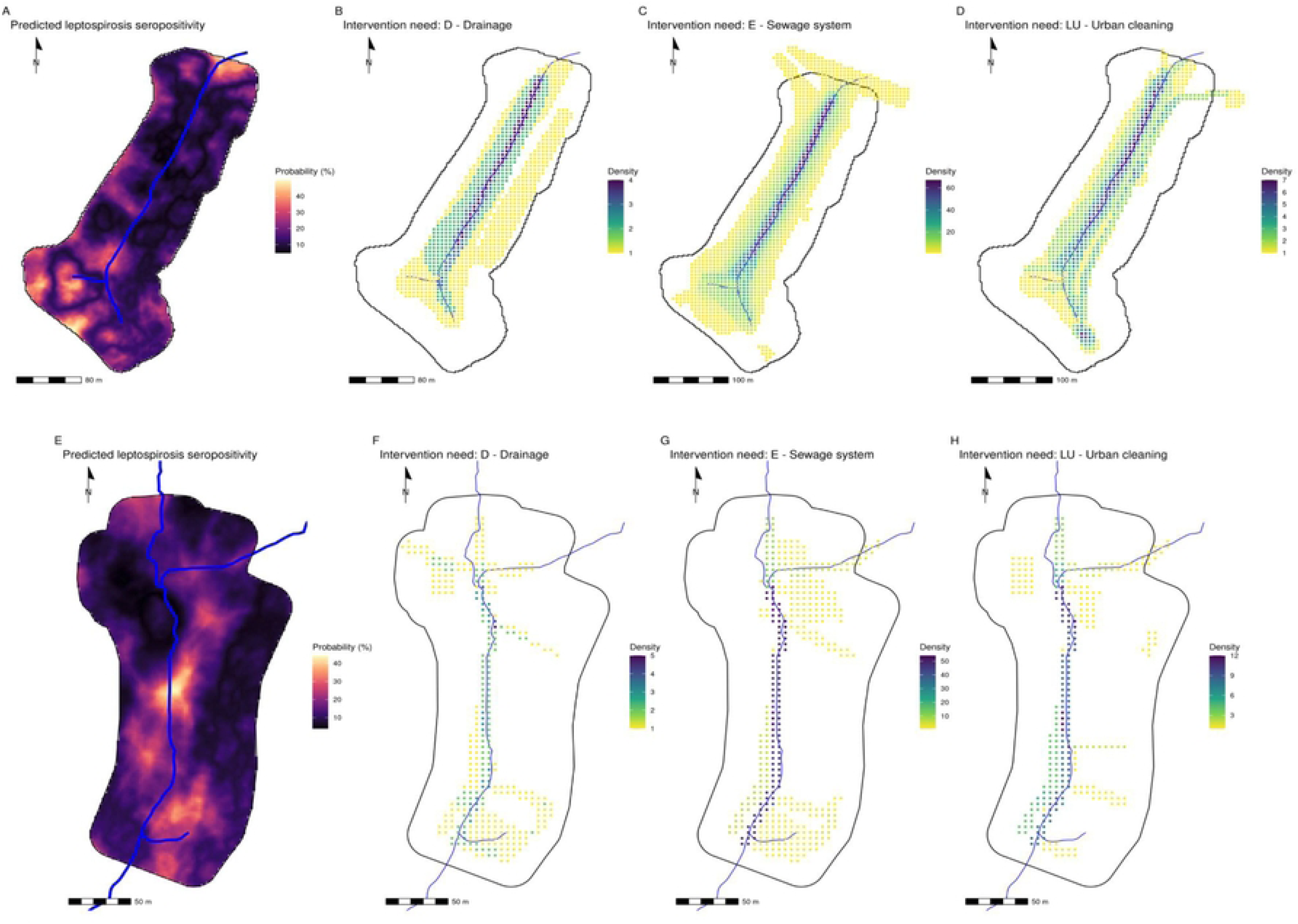
Maps of NS (top row) and JSI/ME (bottom row) showing predicted leptospirosis seropositivity across the study area (A and E) and perceived intervention needs for drainage (B and F), sewage system (C and G) and urban cleaning (D and H). Areas with no dots (white background) mean there were no perceptions from residents. The blue line represents the local river in each study area. The authors created the map in Figure 4 using the results generated from their primary data analysis.

When comparing perceived intervention needs with predicted probability of leptospirosis seropositivity, there was evidence of a negative correlation across most perceptions (Table 4). In NS, for every 1% increase in predicted probability of leptospirosis seropositivity in a 35-year old male, there was a 1 – 2 % reduction in expected density for all perceptions (Sewage: RR = 0.98, 95% CI 0.98 – 0.98, p < 0.001; Urban cleaning: RR = 0.98, 95% CI 0.98 – 0.99, p-value<0.001; Drainage: RR = 0.99, 95% CI 0.99 – 0.99, p = 0.031). In JSI/ME, every 1% increase in predicted probability of leptospirosis seropositivity in a 35-year-old male resulted in a 1% reduction in expected density for perceived need of sewage-related intervention (RR = 0.99, 95% CI 0.99-1.00, p-value<0.001). In contrast, a 1% increase in predicted probability for leptospirosis seropositivity resulted in a 1% increase in expected density for perceived need of urban cleaning (RR = 1.01, 95% CI 1.01 – 1.02, p < 0.001). There was no evidence of a correlation between predicted probability for leptospirosis seropositivity and perceived need for drainage intervention (RR = 1.00, 95% CI 0.99 – 1.01, p = 0.537) (Table 4).

**Table 4.** Estimated effects of predicted probability of leptospiral seropositivity for a 35-year-old male on densities of each type of intervention need, by study area (NS, JSI/ME). Estimates are shown in Rate Ratios.

| Perceived sanitation needs intervention | Communities |  |  |  |  |  |
| --- | --- | --- | --- | --- | --- | --- |
|  | NS* |  |  | JSI/ME** |  |  |
|  | Estimate | 95% CI | p-value | Estimate | 95% CI | p-value |
| Sewage | 0.98 | 0.98, 0.98 | <0.001 | 0.99 | 0.99, 1.00 | <0.001 |
| Urban cleaning | 0.98 | 0.98, 0.99 | <0.001 | 1.01 | 1.01, 1.02 | <0.001 |
| Drainage | 0.99 | 0.99, 0.99 | 0.031 | 1.00 | 0.99, 1.01 | 0.537 |

\*NS: Nova Sussuarana community
\*\*JSI/ME: Jardim Santo Inácio/Mata Escura community

#### Subgroup analysis

In both study areas, there were higher densities of recorded perceptions for women compared to men (supplementary material S4). The relationship between the perceived intervention needs and the predicted probability of leptospirosis seropositivity did not vary when divided into genders.

When looking at the leptospirosis serological status, some differences appeared (supplementary material S5). Firstly, seropositive individuals in NS only identified sewage system interventions as the perceived intervention needs in their community. The relationship between their perception densities and the predicted probability for leptospirosis seropositivity appeared to be non-existent. In JSI/ME, seropositive individuals identified all intervention types as needed in their communities. As in NS, the relationship between the densities of these and the predicted probability for seropositivity appeared to be non-existent. The relationship between the densities of the perceptions for seronegative individuals were the same as described in the full analyses.

## Discussion

This study identified gaps between the perceived basic sanitation needs of residents of peripheral communities and the interventions carried out by the service provider. A relevant feature of this research was the investigation of the perception of these communities in a large urban center in the context of the implementation of simplified sewage systems. In addition, we sought to understand if this perception contributed to exposure to diseases such as leptospirosis, which is associated with inadequate sanitation [29].

Our findings showed that despite sewage being a perceived basic sanitation need, residents also reported urban cleaning and solid waste management require attention, particularly in NS, requiring integrated actions and implementation/installation/construction regarding sewage. In JSI/ME, on the other hand, in addition to the needs related to implementation/installation/construction, maintenance was also an aspect frequently reported by residents. Here, we identified that community perceptions led to intervention recommendations beyond the service provider’s plans, mainly focusing on the sanitary sewage network implementation/installation/construction. Although this was a relevant intervention, a previous study revealed that peripheral areas still lack full functional access to the sanitary sewage network [29]. Our findings integrate a health situation analysis that adopted a collaborative diagnostic approach to analyze the conditions in peripheral urban communities, revealing multiple vulnerability processes [22,38], persistent inequalities in sanitation services [29,39] and high exposure to risk factors for leptospirosis [1,6,9–10].

In Brazil, peripheral urban communities continue experiencing precarious provision of public services, such as sewage and trash collection [40], due to environmental racism [13] and the need for intersectoral actions. Collaborative approaches are needed to address these issues, focusing on identifying problems and disseminating interventions to enact local change based on the perception of those who experience this reality, including public institutions responsible. Even if community collaboration is through questionnaire responses, the potential to capture their perceptions is valid and reveals several processes of vulnerability, including limited participation in decision-making. In Brazil, the most precarious access to sanitation is more frequent among low-income and low-educational level people who self-declare as ethnically black living in the Northeast region [40], such as in the city of Salvador (42)[38], where this study was conducted.

Our findings showed that most residents in NS reported sewer cleaning and sealing, as well as sewer cleaning and piping, as sanitation needs that require integrated intervention. Here, sealing refers to “tamponamento do esgoto” whereas piping means “canalização do esgoto”. The most common implementation/installation/construction intervention was the sanitary sewage network. In JSI/ME, despite the similarity in the implementation/installation/construction dimension, most participants indicated interventions aimed at sewage sealing and piping. Sealing of polluted sewage/river was also a common intervention indication in the communities in our study. This intervention is often associated with inadequate urban intervention projects [41]. However, it is necessary to solve old problems in the study communities, such as the population’s contact with polluted water, which contributes to diseases like leptospirosis [1] and arboviruses [44] and causes discomfort for residents.

Salvador has sealed several rivers, primarily in wealthier neighborhoods or commercial districts, to eliminate odors and mosquitoes and improve landscapes [28–29]. More recently, two remnants of the city’s rivers, the Camarajipe and the Lucaia, were sealed to make the Rapid Transit Bus (BRT) viable [41]. In contrast, peripheral communities face insufficient sewage interventions and sanitation components. While sealing courses of water is in most cases not an appropriated environmental intervention, the population’s right to live without open sewage or contact with polluted river waters, especially during flooding, is a sufficient reason to implement this type of intervention in these contexts. The alternative and more desired option, the remediation and recuperation of courses of water, has been performed in few rivers of urban centers of Brazil, and has not been an available option in peripheral urban communities.

The indications of the types of sewage interventions perceived in the communities also show that, even in contexts where sociodemographic characteristics are homogeneous, as in our study, it is necessary to consider the environmental heterogeneities present in the communities. In our findings, we identified that, in NS there are more households in a situation of housing insecurity located near hillsides, and that do not have paved access to their homes. This scenario, combined with the inadequate disposal of sewage water on hillsides, contributes to landslides, increasing the community’s vulnerabilities and risks [29]. In this study, we identified households in JSI/ME exposed to water insecurity, irregular supply, and a lack of hydrometers. The lack of historical guarantee of this right [31,38] in peripheral urban communities highlights the need for equitable and consistent sanitation and health intervention planning and considering these differences. Thus, planning sanitation and health interventions should be more equitable and consistent with local needs to be more effective in preventing diseases such as leptospirosis, improving the population’s quality of life, and the environmental sustainability of territories [43].

Residents reported that the most common urban cleaning and solid waste management interventions involved removing solid waste in squares and streets. These findings were consistent with a qualitative and collaborative mapping study in four Salvador communities[1,26]. Residents reported addressing waste and sewage problems requires theoretical and practical approaches, such as educational activities and structural interventions. In NS, concerns were mainly related to a container near the community square that accumulates trash, while in JSI/ME, concerns were about trash accumulation at various community points. Trash inside the open sewer was a common concern, indicating insufficient solid waste management practices in poorer areas, such as peripheral urban communities.

We also identified that perceived needs for sanitation in communities correspond to the main risk factors for leptospirosis, as evidenced in previous studies in Salvador [1,9–10]. Contact with sewage water and mud is associated with the risk of human infection by *Leptospira* [10,44]. Furthermore, the most exposed groups are those without access to essential sanitation services and who face greater barriers to adopting preventive practices [45–46]. Progress still needs to be made in addressing risk factors for leptospirosis, such as sewage and trash, in peripheral urban communities. These communities continue having problems that require sanitation intervention in residents’ perception. Furthermore, these problems persist despite the Sustainable Development Goals (SDGs) to address access to drinking water and sanitation by 2030 [49] and the Basic Sanitation Law in Brazil [31]. Given this, we asked: what are the reasons that could explain the maintenance of this situation? What still needs to be done to effectively overcome these sanitation problems, which are risk factors for leptospirosis in these communities? Based on our findings, the current documents and agreements do not adequately guarantee the right to basic services for a dignified and healthy life.

Our analyses showed that residents’ concerns mostly revolve around the central stream in both communities, with most perceptions clustering around this area. When comparing perceived intervention needs with the predicted probability of leptospirosis seropositivity, residents did not indicate that interventions needed to happen in the areas where our model predicted hotspots. Although this could be interpreted as a need for more local education on risks associated with leptospirosis, another interpretation would be that residents were more concerned with the impact of the polluted stream on their daily lives, such as bad smell, mosquito presence, tall vegetation, and frequent blockages and flooding.

The perceived importance of the stream as a matter of health concern, although contrasting with the risk model results, aligns with the ecological importance of the stream as a component of the landscape. Communities built on valleys and floodplains contribute to environmental contamination by pathogens and rat infestation, which are risk factors for leptospirosis [48]. Exposure to *Leptospira* can occur while transiting within the community, not necessarily in the peri-domiciliary environment. The poor environmental health and housing conditions faced in the communities foster rat infestation, allowing the expansion of environmental contamination by *Leptospira* beyond the reach of the stream water [1].

The study has limitations due to its cross-sectional design, which limits inferring causality. Additionally, the study’s focus on residents’ perceptions of sanitation needs with a purely spatial perspective does not provide a comprehensive understanding of the perceptions reported by residents. A qualitative approach could deepen the understanding of these issues. While our study was designed to be representative of the communities, it is possible that community members who were most impacted by basic sanitation problems in the community have been more likely to respond. However, given the high participation rate of over 80% in each community, we expect the effect of this potential bias to be minimal. Another relevant aspect is the possible social and infrastructure differences between the communities, which may have limited the generalization of the results to other peripheral urban communities. This dimension can be influenced by the level of knowledge about basic sanitation and the time they have lived in the communities.

Other important limitations are those associated with our predictive model. Our model predicted the probability of leptospirosis seropositivity using individual serology data. A positive result in this data represents an immune response from being infected with leptospirosis, with or without symptoms. This infection could have been recent or historical. Therefore, it is an imperfect approximation to infection risk. A better measure of objective risk could have been environmental sampling for *Leptospira* bacteria, using either water or soil/mud. The model also shows that an individual lives, or can live, in every 2×2m pixel of the study area. Although we conducted a thorough analysis to identify the best variables to use for the predictive model, there could be other unmeasured variables that can better explain the variation. However, we still believe our model serves as a useful, albeit imperfect, proxy for infection risk.

To our knowledge, this is the first study to analyze sanitation needs based on residents’ perceptions of peripheral urban communities in Salvador, Brazil, considering their spatial distribution and the relationship with the risk of leptospirosis. Our findings are valuable for understanding sanitation needs and their relationship with the risk of leptospirosis in peripheral urban communities and contribute to formulating intersectoral sanitation interventions and public policies aiming to reduce the risk of zoonotic diseases, such as leptospirosis.

This study employs a collaborative mapping methodology to understand local challenges and solutions, focusing on those experiencing them. It values community knowledge, encourages active data collection by community residents, and increases awareness of health risks. This approach can enhance demand for necessary changes to promote local health. However, it acknowledges that communities may face unique challenges, requiring adaptations in study planning. Future research on collaborative mapping may include broader community participation in formulating questionnaires, analyzing data and interpreting results, using other forms of participation that encourage citizenship exercises, educational processes, and reflection on living in the environment of peripheral communities beyond perceptions. Thus, it could address other local needs contributing to the decreased risk of zoonotic diseases, such as leptospirosis, in peripheral urban communities. Combining this methodology with other epidemiological and environmental data would also strengthen the analyses. In addition, new studies could explore different topics related to basic sanitation and the determinants and conditions of health in these communities that impact the increase in cases of zoonotic diseases.

In this study, community insights provided by collaborative mapping could be integrated into ongoing urban planning and sanitation policies through dialogue with government service providers about the results of scientific research like this. Another alternative would be to include participatory methodologies, such as collaborative mapping, in urban planning and sanitation policies to prioritize and identify the locations and types of interventions that must be implemented to ensure they are more socioculturally appropriate and sustainable. Furthermore, in the case of sanitation policies, the sewage intervention model in communities like those in our study has as a guideline the promotion of community participation in the management and maintenance of interventions [21]. Although this is still incipient or absent in practice, it can be strengthened by the results of studies like this.

Finally, in Brazil, an important mechanism to institutionalize community mapping, as carried out in this study, in municipal health and sanitation strategies was the creation of the National Secretariat of Peripheries, linked to the Federal Government, recently published a Popular Mapping Guide [49] that aims to contribute to the strengthening of residents of the peripheries and subsidize the formulation of public policies aimed at these contexts.

## Conclusion

Our findings showed that the prevention of diseases such as leptospirosis in peripheral urban communities requires integrated basic sanitation interventions, encompassing different components and aligned with the local needs perceived by residents. This study aimed to develop a deeper understanding of the health situation of peripheral urban communities in a large urban center based on perceptions of sanitation-related needs and their relationship with diseases such as leptospirosis through a collaborative approach. In addition, we sought to contribute to an evidence base that would assist in the planning of priority sanitation interventions in similar settings characterized by limited resources to make them more effective in preventing zoonotic diseases.

## Authors’ contribution

**Conceptualization:** Fabiana Almerinda G. Palma, Pablo Ruiz Cuenca, Ricardo Lustosa, Federico Costa, Marbrisa N. R. das Virgens, Yeimi Alexandra Alzete Lòpez, Cleber Cremonese, Diogo César de C. Santiago, Ariane Sousa do Carmo, Antonia dos Reis, Gislane de Jesus do Carmo, Andrea Maria Lima, Renata Santos Almeida.

**Data curation:** Fabiana Almerinda G. Palma, Pablo Ruiz Cuenca, Ricardo Lustosa, Marbrisa N. R. das Virgens, Pedro Maciel.

**Formal analysis:** Fabiana Almerinda G. Palma, Pablo Ruiz Cuenca, Ricardo Lustosa, Daiana Santos de Oliveira, Ana Maria N. Silva, Marbrisa N. R. das Virgens, Pedro Maciel, Diogo César de C. Santiago, Ariane Sousa do Carmo, Antonia dos Reis, Gislane de Jesus do Carmo, Andrea Maria Lima, Renata Santos Almeida, Tania Bourouphael.

**Funding acquisition:** Federico Costa, Yeimi Alexandra Alzete Lòpez.

**Investigation:** Fabiana Almerinda G. Palma, Pablo Ruiz Cuenca, Diogo César de C. Santiago, Ariane Sousa do Carmo, Antonia dos Reis, Gislane de Jesus do Carmo, Andrea Maria Lima, Renata Santos Almeida, Lucineide Oliva.

**Methodology:** Fabiana Almerinda G. Palma, Pablo Ruiz Cuenca, Ricardo Lustosa, Daiana Santos de Oliveira, Marbrisa N. R. das Virgens, Pedro Maciel, Ana Maria N. Silva, Juliet O. Santana, Tania Bourouphael.

**Project administration:** Fabiana Almerinda G. Palma, Pablo Ruiz Cuenca, Diogo César de C. Santiago, Ariane Sousa do Carmo, Antonia dos Reis, Gislane de Jesus do Carmo, Andrea Maria Lima, Renata Santos Almeida.

**Resources:** Federico Costa, Yeimi Alexandra Alzete Lòpez, Cleber Cremonese.

**Software:** Fabiana Almerinda G. Palma, Pablo Ruiz Cuenca, Ricardo Lustosa, Marbrisa N. R. das Virgens, Pedro Maciel, Juliet O. Santana.

**Supervision:** Fabiana Almerinda G. Palma, Pablo Ruiz Cuenca, Federico Costa, Emanuele Giorgi, Diogo César de C. Santiago, Ariane Sousa do Carmo, Antonia dos Reis, Gislane de Jesus do Carmo, Andrea Maria Lima, Renata Santos Almeida.

**Validation:** Federico Costa, Yeimi Alexandra Alzete Lòpez, Ricardo Lustosa, Cleber Cremonese, Ana Maria N. Silva, Emanuele Giorgi, Max T. Eyre, Caio G. Zeppelini.

**Visualization:** Fabiana Almerinda G. Palma, Pablo Ruiz Cuenca, Ricardo Lustosa, Marbrisa N. R. das Virgens, Pedro Maciel, Juliet O. Santana, Max T. Eyre.

**Writing – original draft:** Fabiana Almerinda G. Palma, Pablo Ruiz Cuenca, Ariane Sousa do Carmo, Antonia dos Reis, Gislane de Jesus do Carmo, Andrea Maria Lima, Renata Santos Almeida, Lucineide Oliva.

**Writing – review & editing:** Federico Costa, Yeimi Alexandra Alzete Lòpez, Ricardo Lustosa, Cleber Cremonese, Marbrisa N. R. das Virgens, Pedro Maciel, Juliet O. Santana, Ana Maria N. Silva, Daiana Santos de Oliveira, Diogo César de C. Santiago, Tania Bourouphael, Emanuele Giorgi, Max T. Eyre, Caio G. Zeppelini.

## Acknowledgements

We deeply thank the residents and communities of Nova Sussuarana and Jardim Santo Inácio/Mata Escura for participating in this study. We also thank the field research team members for their support and collaborative construction of this research. We thank all the authors and co-authors in the developed this research and the manuscript.

## Availability of data and materials

The datasets used and/or analyzed during the current study cannot be shared publicly because of personal information of participants in the survey. Researchers who wish to access the data can contact the data manager at the Oswaldo Cruz Foundation, Nivison Junior.

## Supporting information

S1 Table. Households’ and peri-domiciliary characteristics of the study area.

S2 Table. Full table of the type of sewage interventions reported by the study population.

S3 Table. Full table of the type of urban cleaning and solid waste management interventions reported by the study population.

S4 Fig. Graphs showing densities for each type of perceived intervention need against predicted probability of leptospirosis seropositivity for a 35-year-old male, separated into gender of participants.

S5 Fig. Graphs showing densities for each type of perceived intervention need against predicted probability of leptospirosis seropositivity for a 35-year-old male, separated into leptospirosis serological status of participants.

S6. Details on leptospirosis seropositivity model

S7. Code analysis_stata_collaborative map.do

S8. Dataset_collaborative map.dta

S9. GAM plots

S10. Univariable estimates table

S11. Model selection table (AIC)

S12. Metadata Orthoimages

